# CO-MORBIDITIES AND ASSOCIATED FACTORS AMONG SEVERELY ACUTELY MALNOURISHED CHILDREN ADMITTED TO PUBLIC HOSPITALS IN NORTH SHOA, ETHIOPIA

**DOI:** 10.64898/2026.03.22.26348202

**Authors:** Mekonnen Kolcha Tumato, Awuraris Hailu Bilchut, Muluken Tessema, Akine Eshete

## Abstract

**Background:** Comorbidity refers to the existence of two or more diseases in the same person. Co-morbidity and severe acute malnutrition have a bidirectional relationship.

**Objective:** To determine the magnitude of comorbidity and associated factors among severely acutely malnourished children who were admitted to public hospitals in north Shoa zone, Ethiopia.

**Methods:** An Institutional-based cross-sectional study design was conducted among children admitted to hospitals from January 20 to May 20, 2024 in the north Shoa zone, Amhara, Ethiopia. Data were collected using a pre-tested, structured questionnaire to collect the necessary information for the study. EPI-info version 7 was used to enter data, and Stata version 16.1 was used to analyze it for both descriptive and analytic analysis. Crude and adjusted logistic regression model were built and a p-value < 0.05 was considered statistically significant.

**Result:** A total of 394 children, 61 had comorbidity of pneumonia and diarrhea. The co-morbidity among severely acutely malnourished children were predicted by food diversity score < 5 groups (AOR = 2.00, 95% CI: 1.09–3.98), single marital status (AOR = 3.00, 95% CI: 1.21–7.65) and very small perceived birth weight (AOR = 7.11, 95% CI: 1.43–35.48).

**Conclusion:** The study revealed a high magnitude of co-morbidity of pneumonia and diarrhea among severely acutely malnourished children who were admitted to selected public hospitals in north shoa zone. Policy makers and health care practitioner establish an integrated intervention to reduce comorbidity.

## Introduction

Pneumonia and diarrhea are the most common diseases associated with severe acute malnutrition (SAM) and pose significant threats to children’s health [1]. In 2019, pneumonia and diarrhea combined accounted for around 5.2 million child deaths around worldwide, they are a common comorbidity associated with severe acute malnutrition [2]. Around the world burden of SAM may therefore beyond the WHO’s startling toll on child survival due to a bidirectional relationship between malnutrition and comorbidity among vulnerable children in low-income and middle-income nations [2, 3].

Despite their having been a reduction in childhood wasting over the last decades, it has been a challenging problem for Africa; particularly the eastern part of the continent [4]. A very recent report revealed that, co-morbid illnesses, primarily infections, pneumonia and diarrhea are the primary cause of children’s difficult SAM issues, particularly in less developed nations like our own [5]. In light of this, the federal minister of health has been managing SAM since 2008 by establishing a therapeutic feeding program, but SAM is still in a hazardous condition and contributes to 23% of child deaths, despite improvements being made in child health nutritional treatments [6].

Despite the efforts of the country on SAM management, it is still a significant problem in Ethiopia and the study area due to pneumonia and diarrhea co-morbidity. Previous researches in the study area were assessed diarrhea and pneumonia with SAM, but no prior research has been done on assessing the simultaneous occurrence of pneumonia and diarrhea with severe acute malnutrition and different contributing factors. This research was tried to identify the commodities of pneumonia and diarrhea with severe acute malnutrition and associated factors. The result of this study will provide a holistic understanding of the co-occurrence of the two conditions. And it helps policymakers and health care providers establish integrated interventions to reduce morbidity and mortality. It provides pertinent information for future planning and for other researchers who are interested in researching the subject; it would act as a stepping stone. It might increase the knowledge acquaintances of the researcher as well as the reader in the area.

## Materials and methods

### Study design

An Institutional based cross-sectional study design was conducted among SAM children who were admitted to public hospitals in the north Shoa zone from April 20 to May 20, 2024.

### Study area and period

The zone is located at 9° 49’ 59.99″ N Latitude and 39° 19’ 60.00″ E Longitude and is bordered on the south and west by the Oromia, on the north by south Wollo, on the northeast by the Oromia zone, and on the east by the Afar region. Based on the 2020 census conducted by the Central Statistical Agency of Ethiopia (CSA), this zone had an estimated total population of 3.5 million. North shoa zone were located 695 km from Bahirdar and 130 kilometers North-East of Addis Ababa. There are two private and eleven public hospitals in north shoa zone; the study was conducted from April 20 to May 20, 2024. This study was conducted in four randomly selected hospitals, which are Mehal Meda Hospital, Debre Brehan Referral Hospitals, shewarobit Hospitals, and Deneba Hospital.

### Study population

All of the children aged from 6-59 months who have co-existence of pneumonia and diarrhea comorbidity among severely acutely malnourished and admitted to four selected governmental hospitals in the North Shoa zone.

### Inclusion criteria

All children aged between 6 to 59 months who were admitted to hospitals with severe acute malnutrition, and have comorbidity from April 20 to May 20, 2024. Severe wasting (marasmus) (W/H < -3 SD or <70%), MUAC <11.5 cm, bilateral edema (Kwashiorkor), family members nearby or guardians, and complete data were used as inclusion criteria.

### Exclusion criteria

Children with a major congenital malformation (i.e., cleft lip/palate, Down’s syndrome, and spinal bifida) or severe neurological impairment (i.e., autism, cerebral palsy, and multiple sclerosis) evaluated on history, examination, or diagnostic tests were excluded.

### Sample size determination and sampling procedure

Sample size was determined using EPI-Info version 7. The single population proportion formula was used by using major determinant variables (male sex, immunization and mother education) from another study. By considering immunization status of children was independent factors since it gave the maximum sample size. From that study, the proportion of immunization status outcome in unexposed group was 19.7%, exposed group was 37.5% and odd ratio was 2.45 [5]. Adding 5% non-response rate, the total sample size was 396.

All children admitted to hospitals with comorbidity of severe acute malnutrition before and during data collection were included as study participants based on their registration documented in registration book. From eleven public hospitals, four were selected by a simple random sampling technique. Then the sample size of the children was calculated and allocated using a proportional allocation formula among four randomly selected hospitals.

And systematic random sampling technique was used for the selection of the study subjects and the caretakers, preferably mothers of children. The registration logbook of under-five inpatient diagnoses was used as a sampling frame to select the study participants (i.e., children aged 6–59 months).

### Data collection methods

Structured questionnaires were employed to collect data. The questionnaire was first prepared in English, and then translated into the local language (Amharic), as the study subjects speak Amharic, and then translated back to check for consistency. Caregivers of children were interviewed, documents were reviewed, and the questionnaire papers were filled out by the interviewers. To assure data quality, a pretest was conducted in Enat Hospital, by taken 5% of the sample size and data were collected using an Amharic questionnaire. Four data collectors and one supervisor were recruited, and one day of training was given by principal investigators on tools, methods of data collection, ethical issues, and the purpose of the study.

## Operational definition

### Severe acute malnutrition

Defined as child diagnosed with severe wasting (marasmus) and bilateral edema (kwashiorkor) and MUAC less than 11.5 centimeters [8].

### Comorbidities

Defined as the co-occurrence of pneumonia and diarrhea comorbidity in the same child at the same time [9].

### Diarrhea

Defined as having three or more watery stools in 24 hours (10).

### Pneumonia

The most common type of lower respiratory infection and the leading infectious cause of mortality in children <5 years (9). And characterized by cough with fast breathing for the age, chest in drawing (11).

### Dietary Diversity

Defines as Consumption of food from at least five groups means that the child has a high likelihood of consuming at least one animal source of food and at least one fruit or vegetable in addition to a staple food [12].

## Data processing and analysis

Data were entered into EPI-Info version 7. Then, data were processed by the STATA 16.1 version for cleaning and analysis. Frequencies and row percentages were obtained from the tabulation. Later, bivariate logistic regression was carried out to assess the effect of individual risk factors on the comorbidity of SAM. Normality test was checked for continuous data and hosmer-Lemeshow was used to assess the fitness of the logistic regression model, which shows that it is fit for the model. When multicollinearity was examined, there was no multicollinearity among the variables (mean VIF = 1.08).

An independent variable with a p value < 0.25 was fitted into a multivariate logistic regression analysis to identify factors significantly associated with the co-morbidity of severely acute malnutrition. Finally, risk factors that were significant in the bivariate logistic regression were incorporated in the multivariate logistic regression. A P value of < 0.05 was considered to be statistically significant.

## Ethical considerations

The study protocol was approved by the institutional review board of Asrat woldeyes health science campus, Debre Brehan University (IRB-202, reference-IRB 01/62/2024). Data were collected after permission was asked from the study subjects following a brief discussion with the caregiver about the purpose and importance of the study for public health. As the study was conducted through no-invasive data collection methods and documents reviewed, no individual patients were subjected to any harm as far as the confidentiality was kept. No personal identifiers were used on data collection. And the recorded data was not accessed to third parts except the principal investigator, and confidentiality was kept.

## Results

The study included 394 children between the ages of 6 to 59 months whose mother or caretakers, delivering a 99.5% response rate. The children’s mean age was 15.40(±0.388) months. In relation to mother-related characteristics, the majority of mothers was married (81.98%), and mothers who had never attended formal schooling (42.39%) (Table1).

**Figure 1:**
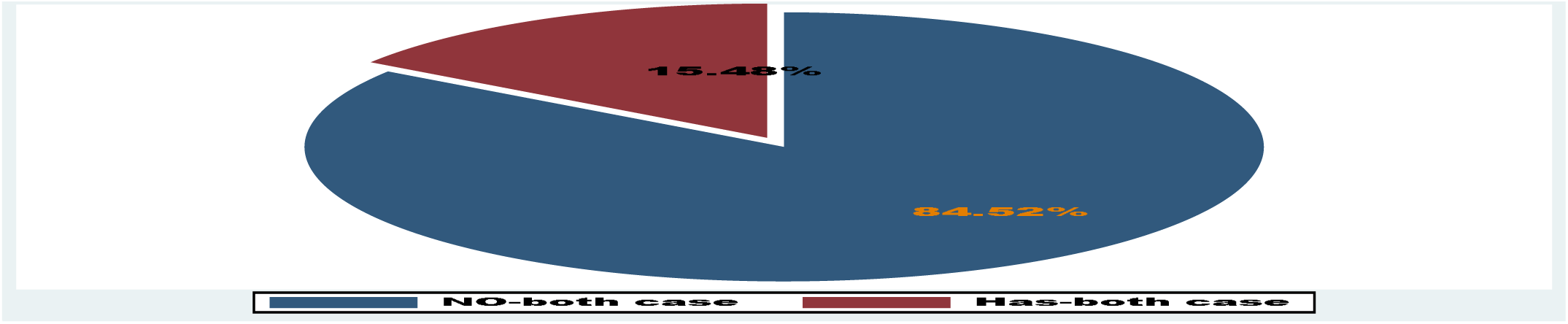
comorbidity of SAM among children who admitted to public hospitals in north shoa zone, Amhara region, Ethiopia, 2024.

**Table 1:**
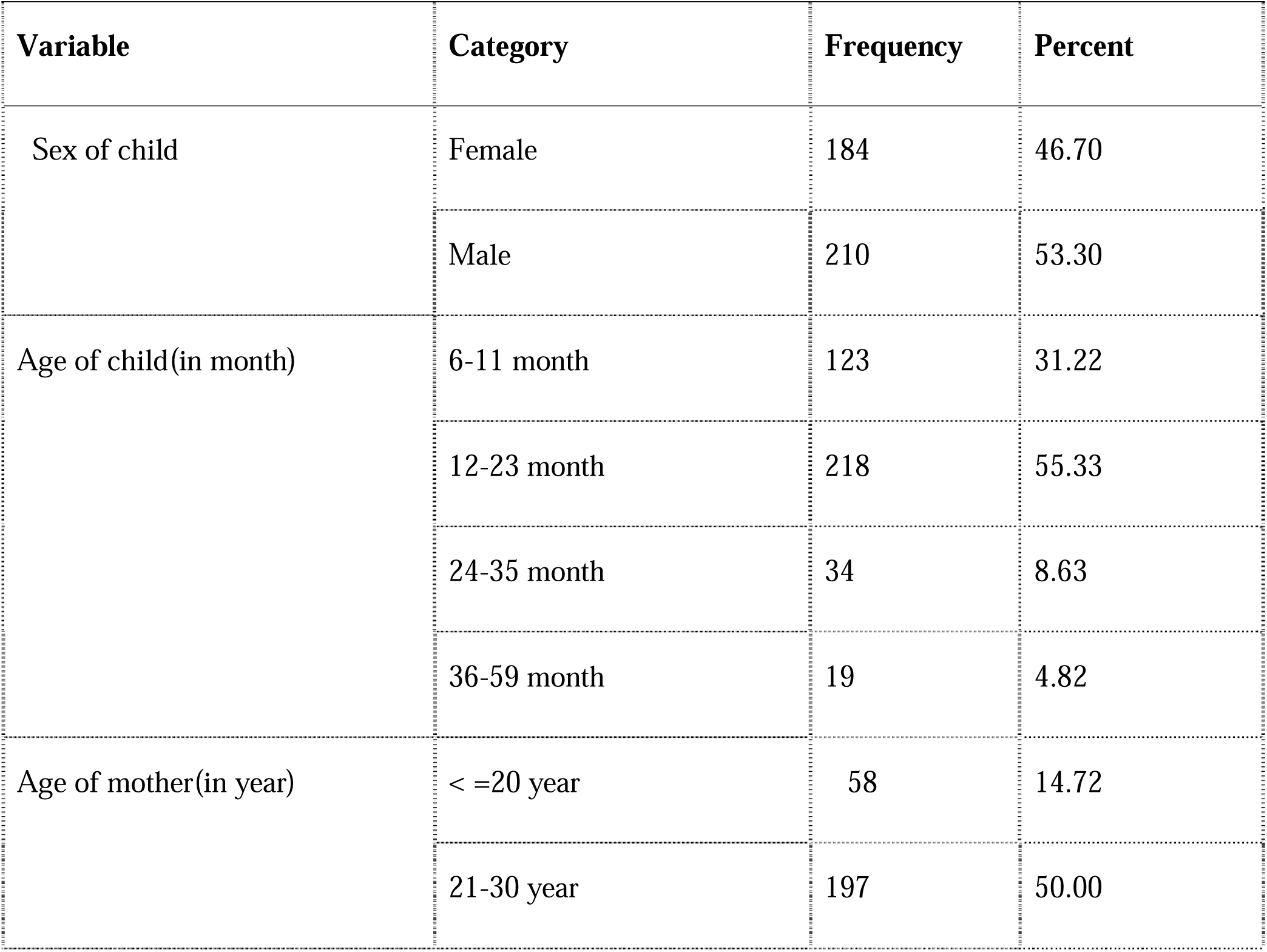
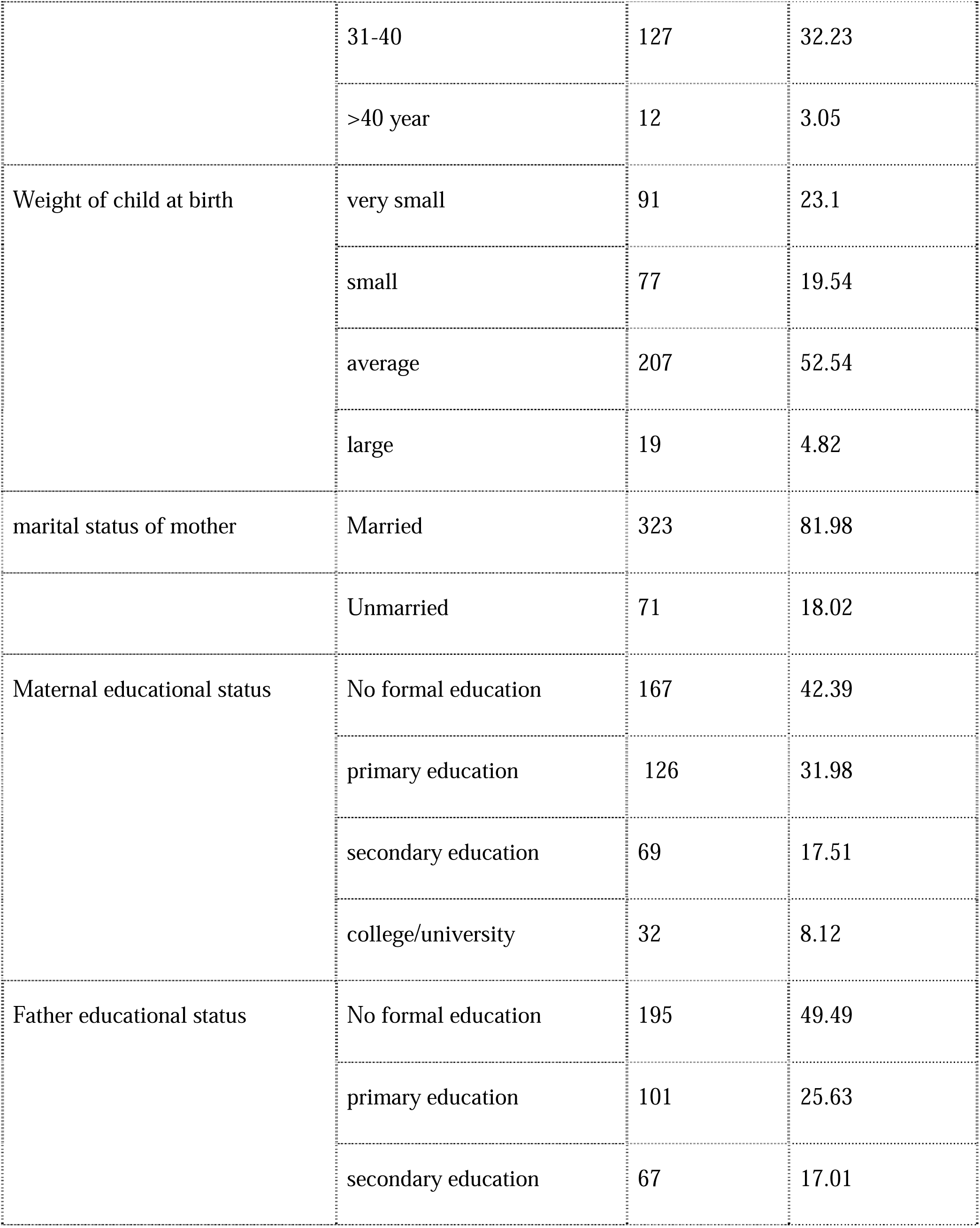
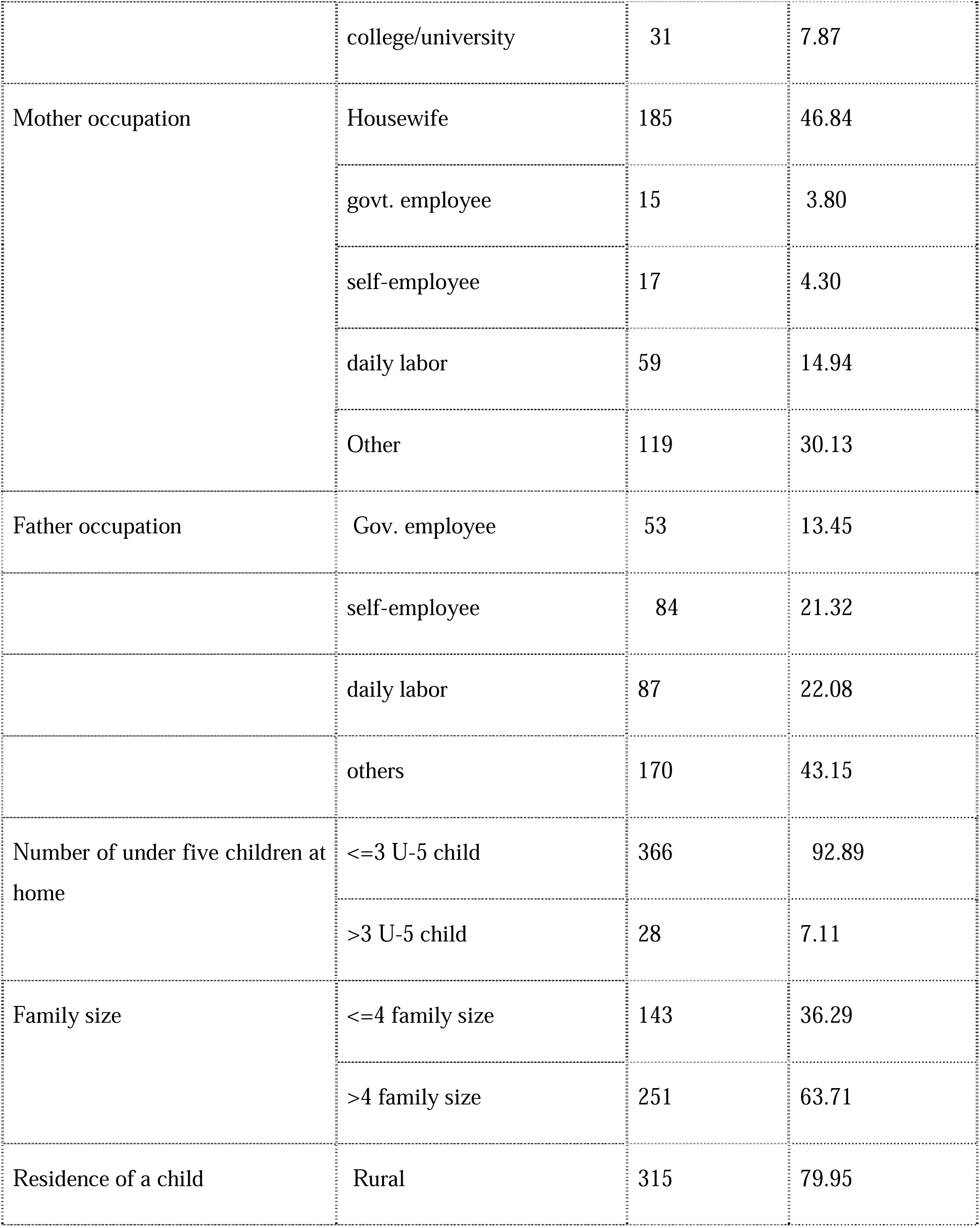
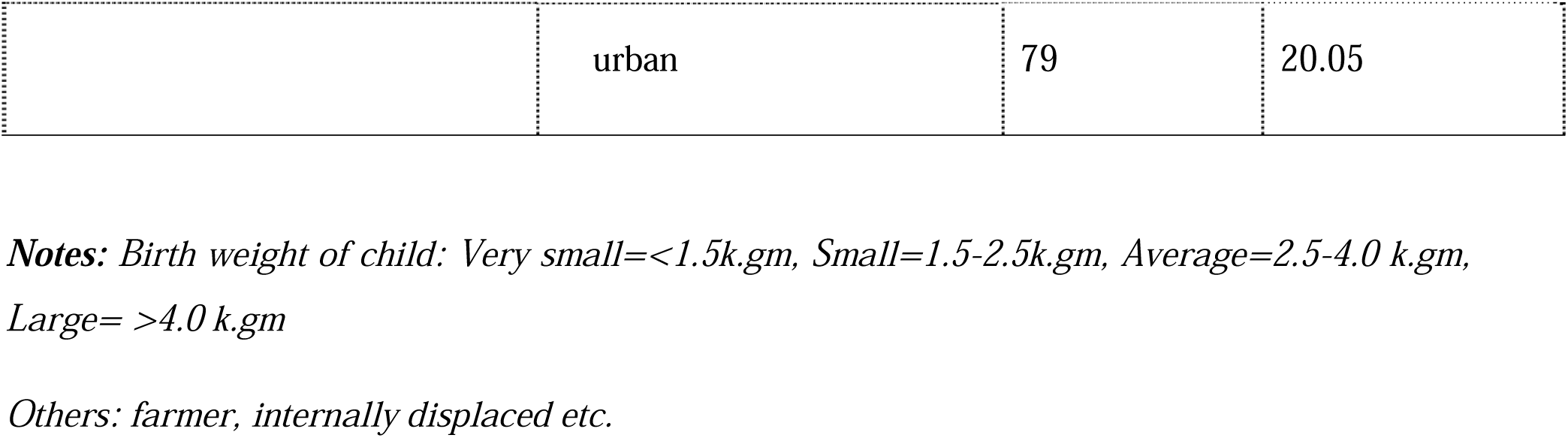
Socio-demographic characteristics of the study participants in north shoa zone, Amhara, Ethiopia, 2024 (n = 394).

| Variable | Category | Frequency | Percent |
| --- | --- | --- | --- |
| Sex of child | Female | 184 | 46.70 |
|  | Male | 210 | 53.30 |
| Age of child(in month) | 6-11 month | 123 | 31.22 |
|  | 12-23 month | 218 | 55.33 |
|  | 24-35 month | 34 | 8.63 |
|  | 36-59 month | 19 | 4.82 |
| Age of mother(in year) | < =20 year | 58 | 14.72 |
|  | 21-30 year | 197 | 50.00 |
|  | 31-40 | 127 | 32.23 |
|  | >40 year | 12 | 3.05 |
| Weight of child at birth | very small | 91 | 23.1 |
|  | small | 77 | 19.54 |
|  | average | 207 | 52.54 |
|  | large | 19 | 4.82 |
| marital status of mother | Married | 323 | 81.98 |
|  | Unmarried | 71 | 18.02 |
| Maternal educational status | No formal education | 167 | 42.39 |
|  | primary education | 126 | 31.98 |
|  | secondary education | 69 | 17.51 |
|  | college/university | 32 | 8.12 |
| Father educational status | No formal education | 195 | 49.49 |
|  | primary education | 101 | 25.63 |
|  | secondary education | 67 | 17.01 |
|  | college/university | 31 | 7.87 |
| Mother occupation | Housewife | 185 | 46.84 |
|  | govt. employee | 15 | 3.80 |
|  | self-employee | 17 | 4.30 |
|  | daily labor | 59 | 14.94 |
|  | Other | 119 | 30.13 |
| Father occupation | Gov. employee | 53 | 13.45 |
|  | self-employee | 84 | 21.32 |
|  | daily labor | 87 | 22.08 |
|  | others | 170 | 43.15 |
| Number of under five children at home | <=3 U-5 child | 366 | 92.89 |
|  | >3 U-5 child | 28 | 7.11 |
| Family size | <=4 family size | 143 | 36.29 |
|  | >4 family size | 251 | 63.71 |
| Residence of a child | Rural | 315 | 79.95 |
|  | urban | 79 | 20.05 |

**Notes:** Birth weight of child: Very small=<1.5k.gm, Small=1.5-2.5k.gm, Average=2.5-4.0 k.gm, Large= >4.0 k.gm
*Others: farmer, internally displaced etc.*

Regarding father education, 195 (49.49%) of the study subjects’ fathers were no formal education. About 43.15% participants father were may be farmer or others and only 13.45% of were governmental workers (Table1).

Considering number of under five children and family size of the study subjects, 366(92.89%) has no more than three under five children at home and 251(63.71%) has greater than four numbers respectively (Table1).

In terms of environmental characteristics, fewer than half (39.39%) of study participants had access to improved water, while more than half (238, or 60.61%) did not. 261(66.24%) of the study participants in this study did not have a latrine. When talking about performing hand washing before food preparation and other study subjects’ activities, 188 (47.72%) have always done so, 206 (52.28%) have done so occasionally, and none have done so never (Table 2).

**Table 2:**
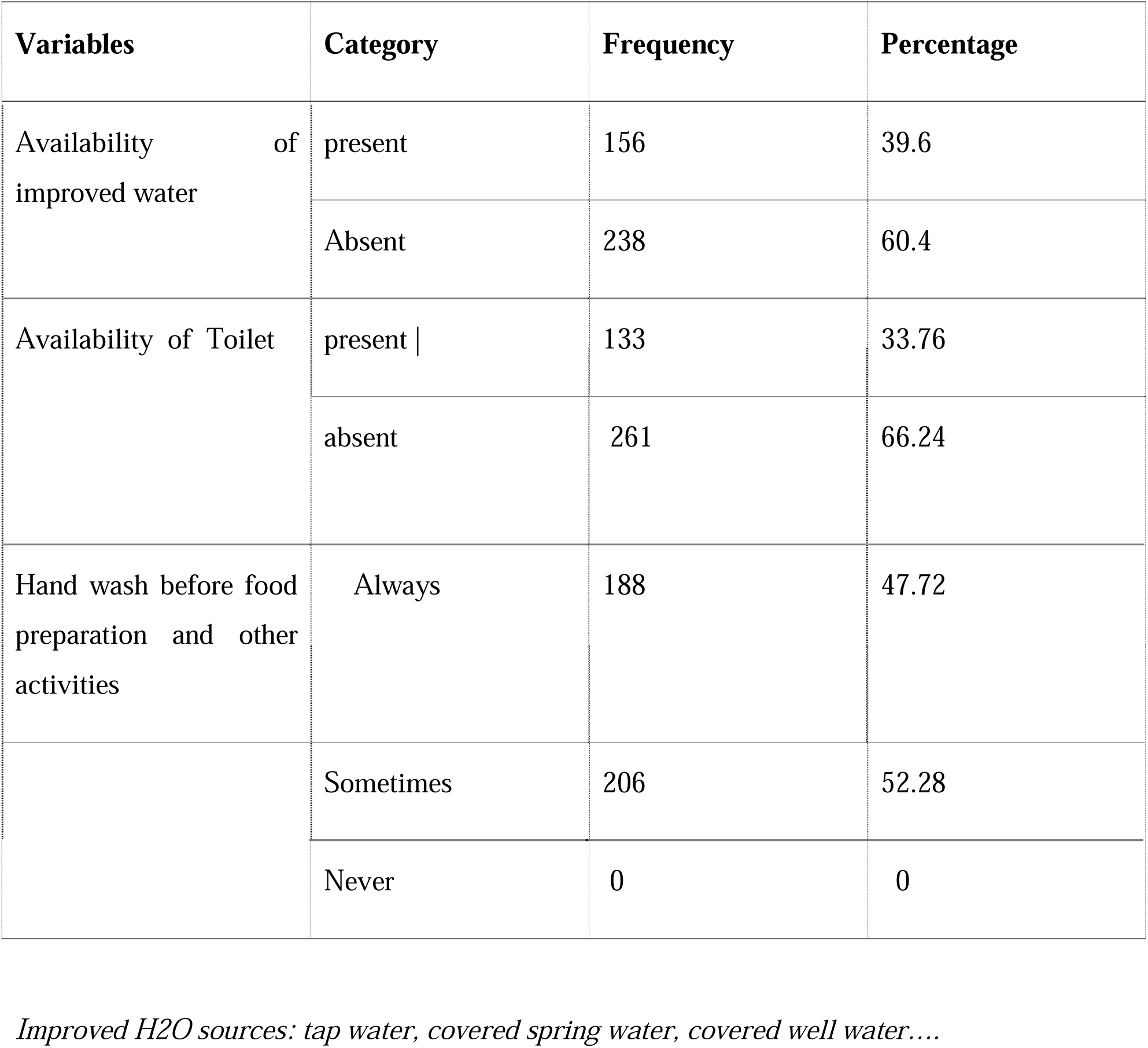
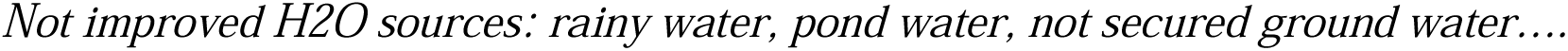
Environmental factors of the study participants in north shoa zone, Amhara, Ethiopia, 2024 (n = 394).

| Variables | Category | Frequency | Percentage |
| --- | --- | --- | --- |
| Availability of improved water | present | 156 | 39.6 |
|  | Absent | 238 | 60.4 |
| Availability of Toilet | present \| | 133 | 33.76 |
|  | absent | 261 | 66.24 |
| Hand wash before food preparation and other activities | Always | 188 | 47.72 |
|  | Sometimes | 206 | 52.28 |
|  | Never | 0 | 0 |

*Improved H2O sources: tap water, covered spring water, covered well water....*
*Not improved H2O sources: rainy water, pond water, not secured ground water....*

81.98% of study participants admitted with single or triple health condition. Of the children who took part in the study, at the time of admission, more than three quarters (295; 75.00%), 50 (12.63%), and 49 (12.37%) had severe marasmus/wasting, edematous/koshakor, and marasmic-kwashkor, respectively. Bilateral edema is one of the criteria used to diagnose an admitted child with severe acute malnutrition. Thirty-five (10.15%) of the three hundred and ninety-four children admitted to the case had grade I edema (+), forty (8.63%) had grade II edema (++), seventeen (4.31%) had grade III edema (+++) and remaining 303(76.90%) were no edema (Table 3).

**Table 3:** Base line clinical and anthropometric information of child in north shoa zone, Amhara, Ethiopia, 2024 (n = 394).

| **Variables** | **Category** | **Frequency** | **Percentage** |
| --- | --- | --- | --- |
| Diagnosis at admission | kwashiorkor | 50 | 12.63 |
|  | wasting/marasmus | 295 | 75.00 |
|  | Marasmic-kwashkor | 49 | 12.37 |
| Presence of edema at Admission | no edema | 303 | 76.90 |
|  | grade 1 edema | 40 | 10.15 |
|  | grade 2 edema | 34 | 8.63 |
|  | grade 3 edema | 17 | 4.31 |
| HIV status | Reactive | 55 | 13.96 |
|  | non-reactive | 153 | 38.83 |
|  | Unknown | 186 | 47.21 |
| Child vaccination status | fully vaccinated | 120 | 30.46 |
|  | partially vaccinated | 154 | 39.09 |
|  | not vaccinated | 103 | 26.14 |
|  | Unknown | 17 | 4.31 |
| single or triple health condition on child | Yes | 323 | 81.98 |
|  | No | 71 | 18.02 |

Regarding the research participants’ HIV status, 153 (38.83%) were non-reactive, 186 (47.21%) had undetermined results, and 55 (13.96%) were reactive (Table 3). In terms of vaccination status, 120 children (or 30.46%) had received all recommended vaccinations, 154 children (or 39.09%) had received some vaccinations, 103 children (or 26.14%) had received none at all, and just 17 children (or 4.31%) had unknown vaccinations (Table 3).

According to feeding practices, 243 (61.87%) of the patients had exclusive breastfeeding for six months, while 151 (38.13%) of the children had no exclusive breastfeeding (Table 4).

**Table 4:** Feeding characteristics of the study participants in north shoa zone, Amhara, Ethiopia, 2024 (n = 394).

| Variables | Category | Frequency | Percentage |
| --- | --- | --- | --- |
| Exclusive breast feed | Yes | 243 | 61.87 |
|  | No | 151 | 38.13 |
| Dietary diversity | >=Five food group | 181 | 45.94 |
|  | < five food group | 213 | 54.06 |
| Age of Starting addition food | < 6 month | 112 | 28.43 |
|  | 6 month | 191 | 48.48 |
|  | 7-12 month | 66 | 16.75 |
|  | >12 month | 25 | 6.35 |
| Means of feeding additional food/drink | Bottle | 104 | 26.40 |
|  | Cup | 28 | 7.11 |
|  | Spoon | 262 | 66.50 |

**Table 5:** Risk factors associated with co-morbidity of pneumonia and diarrhea among severely acutely malnourished children who admitted to governmental hospital in North shoa zone, Amhara, Ethiopia, 2024 (n=394).

| Characteristics | Comorbidity | No-comorbidity | COR (95% CI) | P-value | AOR (95% CI) | P-value |
| --- | --- | --- | --- | --- | --- | --- |
| Marital status of mother |  |  |  |  |  |  |
| Married | 37 | 254 |  | 1 |  |  |
| Single | 10 | 26 | 2.79(1.30-6.01) | 0.008 | 3.00(1.21-7.65) | 0.018 |
| Divorced | 9 | 38 | 1.79(0.84-3.86) | 0.131 | 1.39(0.57-3.34) | 0.467 |
| Widowed | 5 | 15 | 2.31(0.85-5.34) | 0.051 | 2.15(0.68-6.81) | 0.193 |
| Father occupation |  |  |  |  |  |  |
| Gov. Employee | 10 | 43 |  | 1 |  |  |
| Self-employee | 9 | 75 | 0.51(0.19-1.37) | 0.184 | 0.52(0.18-1.48) | 0.219 |
| Daily labor | 17 | 70 | 1.04(0.44-2.49) | 0.922 | 1.17(0.46-2.96) | 0.735 |
| Others | 25 | 145 | 0.74(0.33-1.66) | 0.468 | 0.59(0.24-1.48) | 0.264 |
| Mother occupation |  |  |  |  |  |  |
| House wife | 29 | 122 |  | 1 |  |  |
| Gov. Employee | 1 | 13 | 0.32(0.04-2.57) | 0.286 | 0.28(0.52-4.06) | 0.483 |
| Self-employee | 4 | 35 | 0.48(0.16-1.46) | 0.196 | 0.55(0.16-1.89) | 0.344 |
| Daily labor | 13 | 69 | 0.79(0.39-1.63) | 0.526 | 0.67(0.29-1.51) | 0.332 |
| Others | 14 | 94 | 0.63(0.31-1.25) | 0.186 | 0.73(0.35-1.51) | 0.395 |
| Weight of child at birth |  |  |  |  |  |  |
| Average | 35 | 200 |  | 1 |  |  |
| Large | 15 | 71 | 1.21(0.62-2.34) | 0.578 | 1.19(0.58-2.43) | 0.632 |
| Small | 6 | 58 | 0.59 (0.24-1.47) | 0.260 | 0.63(0.24-1.66) | 0.357 |
| **Very small** | **5** | **4** | **7.14(1.83-27.91)** | **0.005** | **7.11(1.43-35.48)** | **0.017** |

| Dietary diversity |  |  |  |  |  |  |
| --- | --- | --- | --- | --- | --- | --- |
| >5 food group | 20 | 161 |  | 1 |  |  |
| < 5 food group) | 41 | 172 | 1.92(1.08-3.41) | 0.027 | 2.0(1.09-3.98) | 0.027 |
| Number of U-5 children at home |  |  |  |  |  |  |
| Yes | 1 | 27 |  | 1 |  |  |
| No | 60 | 306 | 0.19(0.02-1.42) | 0.105 | 0.16(0.02-1.31) | 0.088 |
| Vaccination status of child |  |  |  |  |  |  |
| Fully vaccinated | 19 | 101 |  | 1 |  |  |
| Partially vaccinated | 20 | 134 | 0.79(0.40-1.56) | 0.504 | 0.71(0.30-0.423) | 0.423 |
| Not vaccinated | 17 | 86 | 1.05(0.51-2.15) | 0.892 | 0.92(0.38-2.19) | 0.844 |
| Unknown | 5 | 12 | 2.21(0.70-7.01) | 0.176 | 1.95(0.48-7.91) | 0.350 |

Regarding dietary diversity, more than half of the study participants had 213(54.06%) taken less than five group, while 181 (45.95%) of the children had feeds more than five food group (Table 4).

Out of study participant, 112(28.43%) were started additional food before the age of 6 months, 191(48.48%) started at the age of 6 months, 66(16.75%) started between the age of 7-12 months and 25 (6.35%) started after 12 months old. Regarding the means of feeding foods other than breast milk, 104(26.40%) are used bottle feeding, 28(7.11%) are used cup and 264(66.50%) are used spoon to feed the child (Table 4).

## Magnitude of co-morbidity of pneumonia and diarrhea among severely acutely malnourished children

In our study, the Double burden of comorbidity of pneumonia and diarrhea among severely acute malnourished was 15.48% (95% CI, 11.89-19.06). According to the multivariate logistic regression analysis, feeding less than five food groups, very small perceived birth weight and single mothers were the only three factors found to be risk factors of co-morbidity of SAM.

Low dietary diversity was found to be significantly associated with the comorbidity of SAM. In the current study, children who feed food group less than five were 2.0 times more likely than counter parts to be affected by a co-morbidity case among severely acutely malnourished children, with a statistically significant association (AOR = 2.0; 95% CI: 1.07-3.91).

The comorbidity of severe acute malnutrition was 3.2 times more likely to affect children of single mothers, according to the mother’s marital status. This indicates a strong association between the variables and a statistically significant association at (AOR=3.2; 95% CI, 1.36-7.57).

Perceived birth weight was shown to be substantially correlated with the comorbidity of severe acute malnutrition in the current study. Very small birth weight of study participants had 7.5 times the chances of having concomitant pneumonia and diarrhea than average perceived birth weight of individuals (AOR=7.5; 95% CI: 1.54-36.62).

## Discussion

In our study the comorbidity of pneumonia and diarrhea among severely acute malnourished children was 15.48%, which is in agreement with the results from Bangladesh (16.3%) [13]. However, this finding is higher than study from Myanmar (3.7%) [14], Ghana(11%) [15]. Similarly, study from Nigeria reported that 9% of under-five children experienced comorbidity [16]. Variations in the socio-demographic characteristics of the study participants, the parents’ socioeconomic status in the communities, or their geographic locations could be the cause of the discrepancy. This high magnitude of 15.48% of children with comorbidity among SAM children has implication for our country far from attaining the sustainable development goal 3(target 3.2) of ending preventable deaths of newborn and children of under-five [17].

The current study also revealed that low dietary diversity (< five food group) was found to be significantly associated with comorbidity of severe acute malnutrition. Children of less than 60 months of age are at greatest risk of malnutrition due to low dietary consumption, infectious diseases and discriminatory food distribution within the family in developing nation [18]. In this sense, in our study the children who feed less than five food group were more affected by the case than children who feed more than five food group. This finding is in line with the Ethiopian demographic and health survey report of 2019 [12] and study from Cameroon [19]. Providing good diversified dietary is important to develop the immune system and preventing infection. So that poor dietary diversity may expose the children to infection due to low immunity [20]. Hence the ongoing conflict in northern Ethiopia since 2021 has probably contributed to the higher likelihood of comorbidity of SAM among children who feed less diversified food. This conflict has resulted in higher food prices, unstable political environments, and job losses, reduced pay, and restricted food availability, which increase the risk of comorbidity in children.

On other hand, the children whose mother was single mom were 3.2 times more likely to be affected by comorbidity than children from married mother. A child’s health and chances of survival are compromised by being raised by single mother. As a result, they are less able than married women to effectively manage the financial aspects of their children’s illnesses [21]. Compared to moms who are married, single mother showed a negative attitude toward the management and prevention of diarrhea. This can be because of their minor financial problems [22]. This finding is consistent with the study conducted from Ethiopia [22], Ivory coast [21], Brazil (23) and Uganda [2].

On other hand, it was observed that very small birth weight was associated with odds of comorbidity than their counterparts. Children with low birth weight are susceptible to infections such as diarrhea and lower respiratory infections [23]. Therefore, Children born with very low birth weight are unable to reach full growth and may therefore be stunted, susceptible to infections and mortality early in life[24].This finding is in line with studies conducted in Ethiopia[20], Mozambique[23] Nigeria[16] and systematic review and meta-analysis from north Africa [25].

## Strength

We are demonstrated existence of co-morbidity of pneumonia and diarrhea with severe acute malnutrition in different hospital in North shoa zone and observed likelihood of increasing child admission with co-morbidity; so this study provide information on risk factors associated with comorbidity of pneumonia and diarrhea among SAM children in North shoa zone.

## Limitations

As this study cross sectional, there was no confirming causality between co-morbidity of SAM and risk factors. May it needs caution in generalization because probability of self-reported information. This study focused on children aged from 6-59 months who experience on both diarrhea and pneumonia. Therefore, the results of this study cannot be generalized to all under five children. Due to this generalized ordinal analysis is better to support this type of study.

## Conclusion

This study revealed high magnitude of comorbidity of severely acutely malnourished children in selected public hospitals. Current study revealed that very low dietary diversity (<5 food group), very small perceived birth weight and single marital status were significantly associated with comorbidity among severely acutely malnourished children.

## Recommendations

Develop strategy for advocating health education for pregnant women, caregivers, and women with children’s of under-five age including knowledge, attitudes, and practices, should be considered, mainly in areas with inadequate dietary diversification for pregnant women and children and the impact of parent separation to reduce the proportion of co-morbidity. And performing routine assessments of their co-morbidity, whether they are suffering from severe acute malnutrition or not, in order to enable early detection, diagnosis, and appropriate treatment.

## Data Availability

All data produced in the present study are available upon reasonable request to the authors

## Author contributions

**Conceptualization:** Mekonnen Kolcha, Awuraris Hailu, Akine Eshete and Muluken Ketema

**Data curation:** Mekonnen kolcha

**Formal analysis:** Mekonnen kolcha, Awuraris Hailu

**Investigation:** Mekonnen Kolcha

**Methodology:** Mekonnen Kolcha, Awuraris Hailu

**Project administration:** Mekonnen Kolcha

**Resources:** Mekonnen Kolcha

**Software:** Mekonnen Kolcha, Awuraris Hailu

**Supervision:** Awuraris Hailu

**Visualization:** Mekonnen Kolcha

**Writing-original draft:** Mekonnen Kolcha

**Writing-review and editing:** Akine Eshete, Meluken tessema and Awuraris Hailu

